# Association between motor task acquisition and hippocampal atrophy across cognitively unimpaired, amnestic Mild Cognitive Impairment, and Alzheimer’s disease individuals

**DOI:** 10.1101/2021.05.31.21258061

**Authors:** Sydney Y. Schaefer, Michael Malek-Ahmadi, Andrew Hooyman, Jace B. King, Kevin Duff

**Author notes:** Corresponding author: Sydney Schaefer, PhD, 501 E. Tyler Mall, MC 9709, Tempe, AZ 85287.

## Abstract

Hippocampal atrophy is a widely used biomarker for Alzheimer’s disease (AD), but the cost, time, and contraindications associated with magnetic resonance imaging (MRI) limit its use. Recent work has shown that a low-cost upper extremity motor task has potential in identifying AD risk. Fifty-four older adults (15 cognitively unimpaired, 24 amnestic Mild Cognitive Impairment, and 15 AD) completed six motor task trials and a structural MRI. Motor task acquisition significantly predicted bilateral hippocampal volume, controlling for age, sex, education, and memory. Thus, this motor task may be an affordable, non-invasive screen for AD risk and progression.

## INTRODUCTION

Hippocampal atrophy is a widely used biomarker for Alzheimer’s disease (AD) stage and progression [1-3]. It is measured using magnetic resonance imaging (MRI), which is cheaper, more widely available, or less invasive than other biomarker testing, such as positron emission tomography and lumbar puncture. However, it still requires extensive equipment, staff, and time, and has a number of contraindications that limit its use among older adults specifically [4]. Factors like claustrophobia or high body mass further restrict MRI use for geriatrics. Older adults are also more likely to move more while in the scanner, affecting scan quality [5]. Furthermore, the need for medical personnel and settings (i.e., hospital) disproportionately discourages under-represented minorities from participating in clinical trials and biomedical research [6, 7] and seeking diagnoses or treatment [8]. Thus, a low-cost, non-invasive, and widely-accessible method to identify hippocampal atrophy in older adults is needed.

Motor behavior may be a potential biomarker that addresses these needs, as complex upper-limb movements have been associated with AD severity [9-11]. Recent work has demonstrated that a rapid, easy-to-administer upper-limb motor task involving adaptive fine motor skill can predict disease progression [12] and is more sensitive to cognitive status than other simple motor assessments [13] while requiring no computer hardware/software. It is feasible for amnestic Mild Cognitive Impairment (aMCI) cohorts to perform [12, 14], and with repeated exposure it can show within-session practice effects (i.e., motor task acquisition) that indicate intact learning ability [14-16] (consistent with [17]). This is in contrast to other motor tasks that require technology (e.g., movement sensors [11], motion capture [10], electromyography [9], or transcranial magnetic stimulation [18]) and often show a ceiling effect. Given the task’s association with disease progression and cognitive status, this short report tested its relationship with hippocampal volume across the AD spectrum (i.e., cognitively unimpaired, aMCI, and mild AD). We hypothesized that motor task acquisition would be related to hippocampal volume, even after controlling for age, sex, education, and memory function.

## METHODS

### Participants

Fifty-four older adults participated in this study, who were a subset of ClinicalTrials.gov NCT03466736 recruited through April 2019. Fifteen were cognitively unimpaired (CU) (mean±SD age = 71.9±4.8 years; 13 females; 17.1±1.8 years of education), 24 were classified with amnestic MCI (aMCI) (mean±SD age = 74.1±5.7 years; 16 females; 15.1±2.7 years of education), and 15 were classified with AD (mean±SD age = 78.6±6.1 years; 7 females; 16.4±2.26 years of education). All participants were White non-Hispanic. Although most aMCI and AD participants were recruited from a cognitive disorders clinic, clinical status was confirmed with the Alzheimer’s Disease Neuroimaging Initiative [19] classification battery, which included the Mini Mental Status Examination [20], Clinical Dementia Rating Scale [21], and the Wechsler Memory Scale–Revised [22] Logical Memory II Paragraph A.

Participants were included if they were ≥65 years of age and had a knowledgeable collateral source available to comment on their cognition and daily functioning. Participants were excluded for medical comorbidities likely to affect cognition (including neurological conditions, current severe depression, substance abuse, and major psychiatric conditions); the inability to complete MRI; the inability to complete cognitive and motor assessments due to inadequate vision, hearing, or manual dexterity; and enrollment in any clinical drug trial related to anti-amyloid agents. Additional exclusion criteria included elevated depression (15-item Geriatric Depression Scale score >5), and moderate or severe dementia (Clinical Dementia Rating score ≥2 or a Mini Mental Status Examination score <20). This study was approved by the University of Utah Institutional Review Board. All participants provided informed consent as self or by proxy prior to enrollment in accord with the Helsinki Declaration of 1975.

Participants underwent extensive neuropsychological assessment; however, only the Delayed Memory Index from the Repeatable Battery for the Assessment of Neuropsychological Status (RBANS, [23]) was examined here for memory function. All subtests were administered and scored as defined in the manual, and normative data from the RBANS manual were used to calculate this Index score as an age-corrected standard score (M = 100, SD = 15) with higher scores indicating better cognition. Mean±SD RBANS Delayed Memory Index scores were 110.5±9.9, 70.1±18.0, and 50.1±9.9 for the CU, aMCI, and AD groups, respectively, consistent with their clinical status.

### Timed motor task

Visual demonstration of the motor task can be viewed on Open Science Framework (https://osf.io/phs57/wiki/Functional_reaching_task/), and its methods have been published previously [12, 14, 24, 25]. This task has also been validated against clinical activities of daily living measures [12] in aMCI patients. To summarize, participants use a standard plastic spoon to acquire two raw kidney beans at a time from a central cup (all cups 9.5cm diameter and 5.8cm deep) to one of three distal cups arranged at a radius of 16 cm at -40°, 0°, and 40° relative to the central cup. Participants used their nondominant hand (to avoid ceiling effects), and started by moving to the cup ipsilateral (same side) of the hand used. They then returned to the central cup to acquire two more beans at a time to transport to the middle cup, then the contralateral cup, and then repeated this sequence four more times for a total of 15 out-and-back movements. Task performance was recorded as trial time (in seconds, via stopwatch); lower values indicate better performance. Movement errors, such as dropping beans mid-reach, were recorded; however, only 1% of all reaches had any errors, and the error rate was similar across groups (p=.70). Participants completed 6 trials of the task. This amount of practice was based on a) previous work demonstrating that cognitively intact older adults typically reach stable performance after 5 trials [16, 26], and b) clinical pragmatism to minimize participant burden (∼5 minutes to administer). Task acquisition was measured as the amount of variability (inter-subject standard deviation) in performance across the practice trials, such that higher standard deviations indicated less task acquisition. Figure 1 illustrates different degrees of acquisition for an individual CU, aMCI, and AD participant, such that the CU participant had better acquisition (less variability) and the AD participant has worse acquisition (more variability) across trials. This illustrates the value of administering more than one trial of the task, allowing for any practice effect. Additional measures of performance included overall mean (averaged across the six trials) and acquisition ‘slope’ (Trial 6 – Trial 1). The measure of acquisition slope is similar to the California Verbal Learning Test learning slope [27], which can differentiate between demented and nondemented older adults [28].

**Figure 1.**
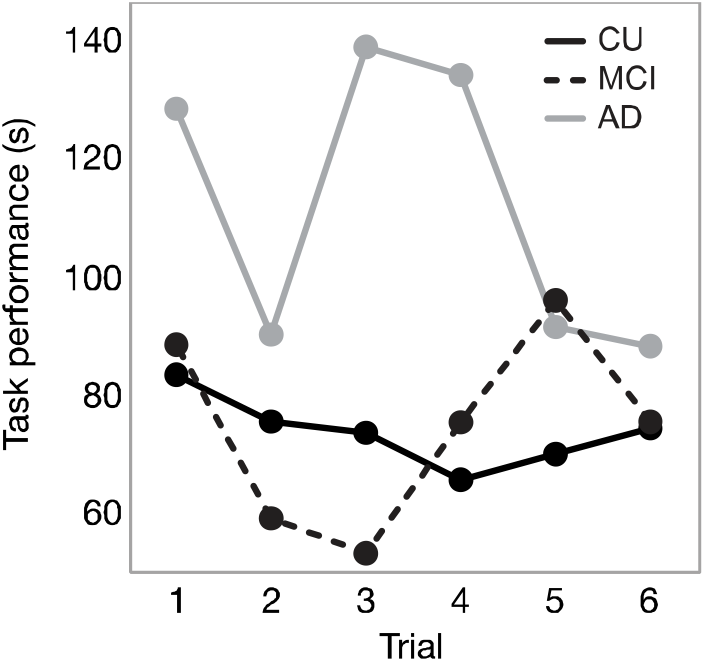
Task performance across six practice trials for a cognitively unimpaired (CU, solid black), aMCI (dashed black), and AD (gray) participant. (Note the difference in variability across the practice trials). Because participants within each clinical status group were not consistently variable across trials (i.e., the AD participants were not all slow on Trial 1, then faster on trial 2, etc.), the average data for each trial for each clinical status group is not shown graphically here; instead, individual participant data are provided to illustrate differences in intrasubject variability.

### MR Imaging Procedure

Acquisition of imaging data was performed at the Utah Center for Advanced Imaging Research (UCAIR) using a 3.0-T Siemens Prisma scanner with a 64-channel head coil. Structural data was acquired using an MP2RAGE pulse sequence (TR=5000, TE=2.93, acquired sagittally, resolution=1 × 1 × 1 mm) to obtain high quality whole-brain 1mm isotropic T1w images with improved signal homogeneity in ∼7 minutes. Structural MRI scans were processed using FreeSurfer image analysis suite v6.0 (http://surfer.nmr.mgh.harvard.edu/). Technical details are described previously [29-31]. Left and right hippocampal volumes were adjusted by estimated total intracranial volume (eTIV, cm^3^) to account for differences in head size, and then summed to yield bilateral hippocampal volumes.

### Statistical analysis

All analyses were performed in R (v3.5.1). Both hippocampal volume and mean task performance were first compared between groups using a one-way ANOVA to determine differences between clinical status. Multivariate linear regression was then conducted to predict bilateral hippocampal volume using participants’ motor task acquisition (i.e., standard deviation) as a predictor while controlling for age, sex, years of education, clinical status, and RBANS Delayed Memory Index score; these factors were included given their known associations with hippocampal volume. The normality assumption for bilateral hippocampal volume was tested using the Shapiro-Wilk test. The motor task variables of acquisition, overall mean, and acquisition slope were separately added to the null regression model (age, sex, years of education, clinical status, and RBANS Delayed Memory Index score) to determine if the contribution of the motor task provided additional predictive value beyond the RBANS Delayed Memory Index score. Akaike’s Information Criteria (AIC) and adjusted R^2^ values from each model were compared.

## RESULTS

One-way ANOVA confirmed a significant effect of clinical status on bilateral hippocampal volumes (F_2,53_ =15.6; p<.0001) (CU = 4.55±0.79 cm^3^, 95%CI [4.11, 4.98]; aMCI = 3.56±0.55 cm^3^, 95%CI [3.33, 3.80]; AD = 3.16±0.89 cm^3^, 95%CI [2.67, 3.65]), consistent with their diagnosis. Furthermore, data showed that this motor task was feasible even for participants with mild AD, even though there was main effect of group on mean task performance (F_2,53_ =5.52; p=.007) with the AD group as the slowest. Mean task performance was not significantly related to hippocampal volume (p=.14), after controlling for age, sex, education, clinical status, and memory score. As shown in the individual participant data in Figure 1, the AD participant was not only slower but also had greater variability across trials (i.e., less task acquisition). Across participants, this variability across trials was sensitive to hippocampal volume, given that regression analyses revealed that motor task acquisition (measured as standard deviation of performance across the 6 practice trials) was a significant predictor of bilateral hippocampal volume (β=-.03; 95%CI [-.07, -.0009]; p=.04), even when controlling for age (p=.46), sex (p=.50), education (p=.33), clinical status (p=0.25), and memory score (β=.01; 95%CI [-.003, .02]; p=.11). The full model yielded an adjusted R^2^ =.36 (F_4,47_=6.1; p<.0001). Comparison of the linear regression models demonstrated that adding either the motor acquisition or mean motor task performance variable to the null model yielded incremental improvements in predicting hippocampal volume, while adding acquisition slope increased the percent variance explained in hippocampal volume by 9% compared to the RBANS Delayed Memory Index (Table 1).

**Table 1.** Linear regression results for models with each motor task variable as a predictor of hippocampal volume, compared to the null model including RBANS Delayed Memory Index. Age, sex, years of education, and clinical status are controlled for in each model.

| Model | $\beta$ | 95% CI | p-value | $R^2$ | AIC |
| --- | --- | --- | --- | --- | --- |
| RBANS Delayed Memory Index | 0.009 | (-0.005, 0.02) | 0.21 | 0.32 | 128 |
| Motor task acquisition (SD) | -0.03 | (-0.06, -0.009) | 0.04 | 0.36 | 125 |
| Mean motor task performance | -0.008 | (-0.02, 0.003) | 0.14 | 0.34 | 127 |
| Acquisition slope (Trial 6-Trial 1) | 0.02 | (0.004, 0.03) | 0.007 | 0.41 | 121 |

## DISCUSSION

This study tested the relationship between bilateral hippocampal volume and acquisition of a motor task in cognitively unimpaired, aMCI, and mild AD older adults. Results showed that even after controlling for age, gender, education, clinical status, and memory, motor task acquisition was still a significant predictor of hippocampal volume, with worse task acquisition (i.e. more variable performance) being associated with lower hippocampal volume. This suggests that motor practice effects may better indicate hippocampal atrophy even after controlling for other clinical factors, which is particularly relevant for cases of MRI contraindication.

Although several complex upper-limb tasks have been shown to be sensitive to disease severity [9, 11], this is among the first to associate motor behavior with an AD biomarker. This work highlights the value of evaluating multiple trials of a motor task, rather than a “one-and-done” approach in which a single attempt could mask relevant differences. This is consistent with extensive work showing the clinical utility of cognitive practice effects [32-36]. Furthermore, these findings are consistent with behavioral data linking practice effects on this motor task with visuospatial scores [16, 37, 38], suggesting a potential mechanism underlying the relationship to hippocampal volume shown here. Future research is needed, however, to examine whether declines in motor acquisition track with hippocampal atrophy (or other biomarkers) over time.

We acknowledge the high education levels and lack of racial/ethnic diversity within the relatively small sample. These limitations warrant future research in larger and diverse cohorts to better estimate the potential of this task as an affordable enrichment strategy for AD clinical trials. We also acknowledge that this study does not directly compare this motor task to other existing motor tasks (e.g., grip dynamometry, 10-Meter Walk Test), although we have previously shown that the motor task presented here is more sensitive to disease severity [24].

In addition to the growing evidence for this motor task as a relevant measure for AD, it should be highlighted that only ∼5 minutes are needed to administer several trials, and the apparatus costs <$10 to fabricate from household items, thereby potentially improving detection of hippocampal atrophy with virtually no additional time or cost. It is also extremely portable, making it easy to administer outside of a clinical setting (e.g., at a community center or at home). Eliminating the need for medical staff/settings has the potential to better serve under-represented minorities. Future studies will test the reliability of administering this motor task in various settings across a more diverse sample.

## Supporting information

STROBE checklist

## Data Availability

Data are unavailable at this time.

